# Protocol for the Enhanced Management of Multimorbid Patients with Chronic Pulmonary Diseases: Role of Indoor Air Quality

**DOI:** 10.1101/2024.05.17.24307036

**Authors:** Alba Gómez-López, Ebymar Arismendi, Isaac Cano, Ramón Farré, María Fígols, Carme Hernández, Antonio Montilla-Ibarra, Núria Sánchez-Ruano, Benigno Sánchez, Antoni Sisó-Almirall, Marta Sorribes, Emili Vela, Jordi Piera-Jiménez, Jaume Benavent, Jose Fermoso, Josep Roca, Rubèn González-Colom

## Abstract

**Introduction:** Reducing unplanned hospital admissions in chronic patients at risk is a key area for action due to the high healthcare and societal burden of the phenomenon. The inconclusive results of preventive strategies in patients with chronic respiratory disorders and comorbidities are explainable by multifactorial but actionable factors.

The current protocol (January 2024 to December 2025) relies on the hypothesis that intertwined actions in four dimensions: i) management change, ii) personalisation of the interventions based on early detection/treatment of acute episodes and enhanced management of comorbidities, iii) mature digital support, and iv) comprehensive assessment, can effectively overcome most of the limitations shown by previous preventive strategies. Accordingly, the main objective is to implement a novel integrated care preventive service for enhanced management of these patients, as well as to evaluate its potential for value generation.

**Methods and analysis:** At the end of 2024, the specifics of the novel service will be defined through the articulation of its four main components: i) Enhanced lung function testing through oscillometry, ii) Continuous monitoring of indoor air quality as a potential triggering factor, iii) Digital support with an adaptive case management approach, and iv) Predictive modelling for early identification and management of exacerbations. During 2025, the novel service will be assessed using a Quintuple Aim approach. Moreover, the Consolidated Framework for Implementation Research will be applied to assess the implementation. The service components will be articulated through four sequential six-months Plan-Do-Study-Act cycles. Each cycle involves a targeted co-creation process following a mixed-methods approach with the active participation of patients, health professionals, managers, and digital experts.

**Ethics and dissemination:** The Ethics Committee for Human Research at Hospital Clinic de Barcelona approved the protocol on June 29, 2023 (HCB/2023/0126). Before any procedure, all patients in the study must sign an informed consent form.

**Registration:** NCT06421402.

**STRENGTHS AND LIMITATIONS:**

- The articulation of the four service components, identified in a previous co-creation process, ensures a comprehensive approach to the novel integrated care service.
- Innovative features of the digital support to the service provided by the Health Circuit platform, based on an Adaptive Case Management orientation, should contribute to overcoming current limitations in digital health transformation.
- Using a Plan-Do-Study-Act strategy, with targeted objectives for each co-creation cycle, will ensure the maturity of the service design and evaluation processes throughout the protocol’s execution.
- The comprehensive evaluation of the novel service, including a Quintuple Aim approach and assessment of the deployment process using the Consolidated Framework for Implementation Research, will contribute to overcoming the efficacy-effectiveness gap seen in this type of integrated care service.
- A potential constraint is that only some patients within the cohort may qualify as future candidates for the innovative service in an actual working environment. Nonetheless, the established protocol is designed to effectively discern and identify the specific patient profiles who stand to derive notable benefits from these interventions.

## 1. INTRODUCTION

Unplanned hospital admissions in multimorbid patients with a high risk of acute events generate significant healthcare and societal dysfunctions, as well as high financial burden^1,2^. Early treatment of exacerbations and enhanced management of comorbidities^3,4^ are the two major actionable factors to avoid unplanned hospital admissions. However, the value generation of preventive strategies in patients with chronic respiratory diseases and comorbidities is inconclusive^5,6^. Such lack of results can be attributed to several factors, namely: heterogeneities of patients with asthma or chronic obstructive pulmonary disease (COPD)^7^, dominant role of co-morbid conditions in the acute episodes^8^, unawareness of potentially relevant triggering factors^9–11^, diversity of the interventions and/or suboptimal description of the protocols leading to poor comparability^12,13^, among other aspects. Overall, there is a clear consensus on the need to explore the potential of clinical processes to efficiently prevent unplanned hospital admissions within an integrated care approach^14,15^ in chronic respiratory patients with multimorbidity.

To this end, a recent co-creation process^16^ was undertaken as part of a large international project^17^ to scale up integrated care services across twenty-one European Union (EU) countries/regions. This process delved into the challenge of preventing unplanned hospital admissions in high-risk candidates with multimorbidity, in whom the severity of the primary disease exerts significant influence^18^. Key lessons learned were the need for specific actions on four dimensions to achieve: i) early identification of the acute episodes and adjustment of organisational aspects of management, ii) personalisation of interventions, iii) mature digital support to the clinical services, and iv) enhanced applicability of the assessment of the service deployment in real-world scenarios.

The current protocol, emerging from the co-creation process, will be executed over two years (from January 2024 to December 2025) to explore the hypothesis that intertwined actions in the above mentioned four dimensions can effectively overcome most of the limitations shown by previous strategies aiming at preventing unplanned hospital admissions in chronic respiratory patients with a high risk of severe acute events. Accordingly, the study’s main objective is to implement a novel integrated care preventive service for enhanced management of these patients and to evaluate its potential for value generation in a real-world scenario.

At the end of the first year (2024), the aim is to define the specifics of the novel service through the articulation of its four main components: i) Enhanced lung function testing through oscillometry^19^, ii) Continuous monitoring of indoor air quality (IAQ) as potential triggering factor^9–11^, iii) Digital support with an adaptive case management approach^20^, and iv) Predictive modelling for early identification and management of exacerbations.

During the second year (2025), the protocol aims to assess the novel service using a Quintuple Aim^21^ approach. It will evaluate the implementation process using a highly applicable version of the Consolidated Framework for Implementation Research (CFIR)^22^. The latter seeks to identify facilitators/barriers to successful service transferability to other sites.

In summary, the current study aims to explore the potential for value generation of the approach to be further scaled up in real-world settings. The novel service, tested in the protocol, will be addressed to community-based chronic patients showing a high risk of hospital admissions, as well as to target patients during transitional care after hospital discharge.

### 1.1 The context

The protocol has been built up under the umbrella of the EU project “Knowledge for improving indoor AIR quality and HEALTH” (K-HEALTHinAIR, 2022-2026)^23^. The primary goal of this EU project is to enhance understanding regarding the impact of chemical and biological IAQ on human health. Moreover, it aims to devise cost-effective methods for more precise monitoring and improvement of IAQ. This will be achieved by analysing data gathered from ten real-life scenarios across Europe, emphasising public health surveillance, and focusing mainly on vulnerable demographics such as high-risk patients, older adults, and children. A detailed description of the K-HEALTHinAIR can be found in the **Supplementary Material – Appendix 1**.

Within this framework, the research team at the Integrated Health District of Barcelona-Esquerra^24^ (AISBE) is conducting a follow-up study of a cohort of multimorbid patients with obstructive lung disorders to assess the relationships between IAQ and health status. This unique setup provides an optimal environment for testing and refining integrated care preventive interventions to reduce unplanned hospitalisations.

## 2. METHODS AND ANALYSIS

Figure 1 depicts the general structure of the current protocol. It is conceived as a two-phase prospective follow-up of 200 outpatients from January 1, 2024, to December 31, 2025.

**Figure 1.**
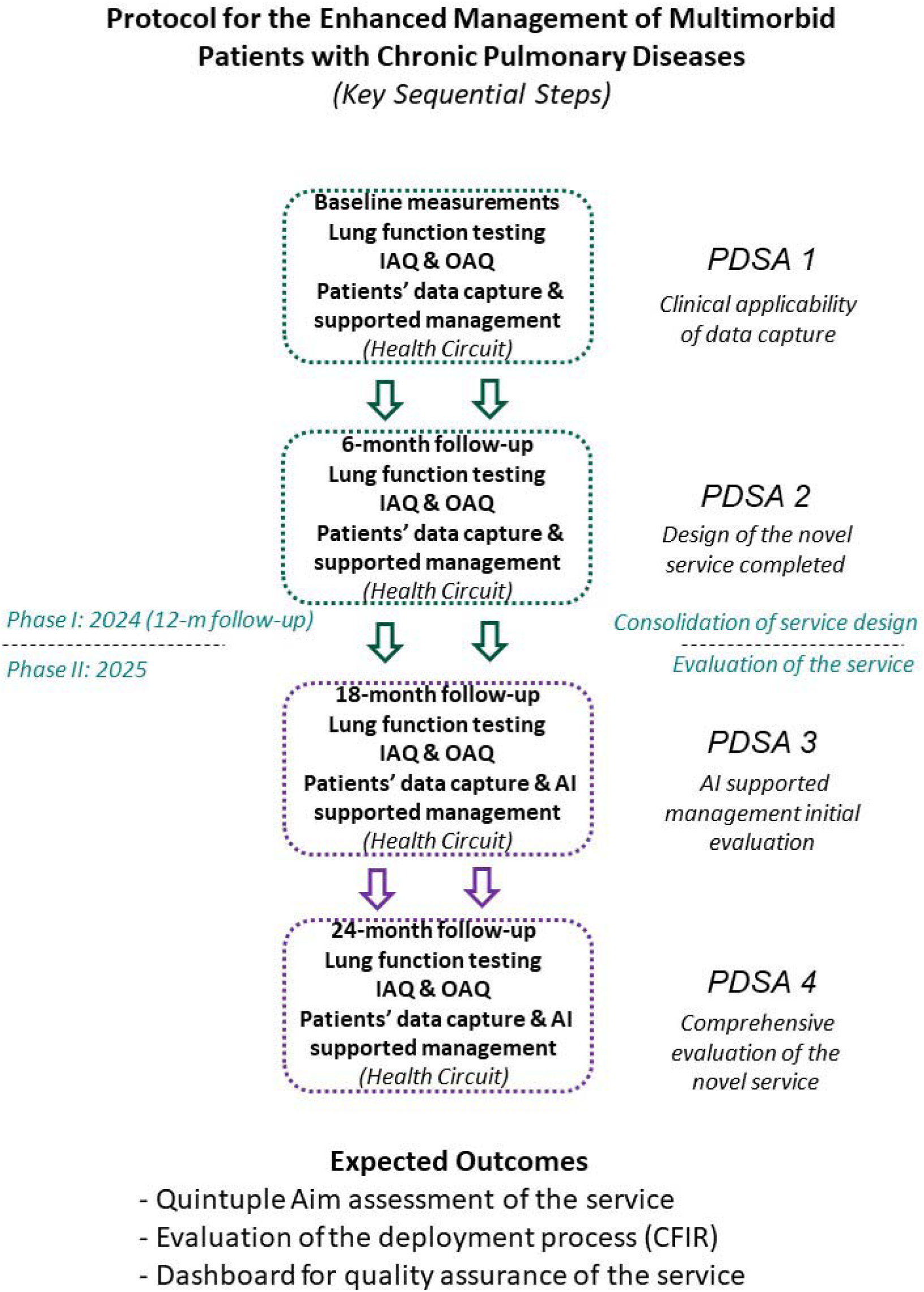
Study protocol flowchart. The four main components of the service are designed and assessed at the end of the first year. The second year, 2025, addresses evaluating the novel service following a Quintuple Aim^21^ approach and assessing the implementation process using the Comprehensive Framework for Implementation Research (CFIR)^22^. Throughout the protocol, four Plan-Do-Study-Act (PDSA)^25^ co-creation cycles, six months duration each, will be conducted with the participation of patients, health professionals, managers, and digital experts. See text for detailed descriptions. IAQ and OAQ stand for Indoor and Outdoor Air Quality respectively; AI stands for Artificial Intelligence.

Phase I of the protocol, running during the entire 2024, has been designed to consolidate the characteristics of the novel service at the end of the first year, including the articulated execution of its four main components. Likewise, the main objective of Phase II, running during the entire 2025, is threefold: i) Assessment of the service outcomes, ii) Identification of the profiles of patients’ candidates for preventive interventions, and iii) Evaluation of the implementation process.

The articulation of the different service components defined in the protocol will be achieved through four sequential Plan-Do-Study-Act (PDSA) cycles^25^ of six months duration each (Figure 1), following the protocol used in^17^. Each PDSA cycle involves a targeted co-creation process following a mixed-methods approach, including quantitative and qualitative methodologies, with the active participation of patients, health professionals, managers, and digital experts.

The expected achievements at the end of the PDSA cycles are as follows: i) PDSA-1: Selection of clinically applicable patients’ data capture for the different dimensions depicted in **Table 1**; ii) PDSA-2: Completion of the design of the novel service; iii) PDSA-3: First assessment of the predictive modelling for early detection of exacerbations facilitating AI supported management of the patients; and iv) PDSA-4: Comprehensive assessment of the novel service. The details of the PDSA method, the templates used to monitor each part of the cycle, and the “Plan” for the initial cycle can be found in **Supplementary Material – Appendix 3.**

**Table 1.** Main data domains considered in the baseline assessment and during the two-year follow-up of the cohort (n=200)

| K-Health in Air project |  |  |
| --- | --- | --- |
| **Standardised questionnaires** | Baseline & every six months | **PROMs:** ICHOM Adult Set<sup>31</sup>, which includes PROMIS 10<sup>32</sup>, WHO 5<sup>33</sup> and WHO-DAS 12<sup>34</sup> questionnaires.<br>**PREMs:** PaRIS<sup>35</sup> survey.<br>**Measurements:** Systemic arterial pressure and pulse oximetry. |
| **Lung Function Testing** | Baseline & every six months | **Forced Spirometry:** FVC and FEV1.<br>**Oscillometry:** Impedance, Resistance and Reactance. |
| **EMRs & Registry** | Baseline & every twelve months | **EMRs:** from the hospital, primary care and regional shared data.<br>**Registry data:** from the CHSS<sup>26</sup>. |
| **Indoor Air Quality (IAQ)** | Continuous monitoring | **MICA-INBIOT system**<sup>36</sup>: temperature, humidity, CO<sub>2</sub>, total VOCs, Formaldehyde; and PM 1/2.5/4/10. |
| **Outdoor Air Quality (OAQ)** | Continuous monitoring | **Aeris Weather platform**<sup>37</sup>: NO, NO<sub>2</sub>, NO<sub>x</sub>, SO<sub>2</sub>, SO<sub>3</sub>, CO, and PM10. |
| Enhanced patient management |  |  |
| **Disease-specific questionnaires** | Baseline & every six months | **COPD:** CAT<sup>38</sup>; mMRC Dyspnea scale<sup>39</sup>.<br>**Asthma:** ACT<sup>40</sup>; TAI-10<sup>41</sup>; mini AQLQ<sup>42</sup>; and SNOT-22<sup>43</sup>. |
| **Physiological variables** | Continuous monitoring | **Beat one watch**<sup>44</sup>: steps walked and heart rate. HRV, sleep structure, stress level as derived variables. |
| **Communication channel** | As needed | **Health Circuit**<sup>20</sup>: chat, short questionnaires (Likert scale) |
| **Personalized follow-up plan** | Baseline & as needed | **Health Circuit**<sup>20</sup>: follow-up of the personalised action plan agreed with the patient & reference doctor (visit 3 at baseline). |
| **Characteristics of exacerbations** | As needed | **Health Circuit**<sup>20</sup>: home-based oscillometry and daily disease-specific questionnaire during the acute episode. |

*List of acronyms sorted alphabetically:* ACT (Asthma Control Test), AQLQ (Asthma Quality of Life Questionnaire), CAT (COPD Assessment Test), CHSS (Catalan Health Surveillance System), FEV1 (Forced Expiratory Volume in 1 second), FVC (Forced Vital Capacity), ICHOM (International Consortium for Health Outcomes Management), K-HEALTHinAIR (EU Project "Knowledge for improving indoor AIR quality and HEALTH"), mMRC (modified Medical Research Council), PaRIS (Patient-Reported Indicator Survey), PM (Particulate Matter), PROMIS (Patient-Reported Outcomes Measurement Information System), PROMs (Patient Reported Outcome Measures), PREMs (Patient Reported Experience Measures), SNOT (Sino-Nasal Outcome Test), TAI (Test of Adherence to Inhalers), VOC (Volatile Organic Compound), WHO (World Health Organization).

***Patients’ enrolment, from November 2023 to the end of June 2024:*** The recruitment efforts for 80% of the study group spanned four primary care facilities from AISBE, each with a catchment population of approximately 20k citizens. The first step was elaborating the dataset of potential candidates using registry information from the Catalan Health Surveillance System (CHSS)^26^, allowing the identification of the corresponding primary care physicians.

The primary care physicians’ engagement in the study facilitated contact with the patients, exploring their willingness to participate and keeping them within their healthcare team during the follow-up period. An initial phone visit of the nurse case manager, on behalf of the primary care team, is used to introduce the project and set the first face-to-face visit at the patient’s home.

The remaining 20% of the cohort includes patients with severe asthma (GINA^27,28^ step 5) recruited from the outpatient of Severe Asthma Unit of the Pulmonology Department of Hospital Clinic de Barcelona (HCB) following similar steps.

***Inclusion criteria:*** i) adults <85 years, ii) diagnosis of chronic obstructive pulmonary conditions (i.e. COPD or severe asthma), iii) showing a high burden of comorbidities, above percentile 80 of the regional risk stratification pyramid, assessed by Adjusted Morbidity Groups (AMG) scoring^29,30^, and iv) living in AISBE, except for patients with asthma resistant to standard therapy that due to the scarcity of cases the catching area has been extended to any the entire city of Barcelona.

The AMG is a morbidity grouper designed to capture patients’ disease burden by considering the quantity and complexity of concurrent conditions. It utilises disease-specific weights derived from statistical analyses incorporating mortality rates and healthcare resource utilisation.

***Exclusion criteria:*** i) patients with dementia or unable for independent daily life activities.

***Data acquisition:* Table 1** indicates the data collection strategy considered in the current study. The top section, titled *“K-Health in Air project”,* describes the categories of information collected as background data within the EU project. In contrast, the bottom part, named *“Enhanced patient management”,* indicates the data categories considered for characterisation and early detection of exacerbations in these patients.

The baseline evaluation and follow-up of the participants includes an assessment of their digital literacy such that the study protocol considers the following three modalities of patients’ participation in terms of technological support, namely: i) phone-based only, ii) phone-based and passive use of the Beat One watch sensor^44^, and iii) digital support for patients and health professionals provided by the Health Circuit platform^20^. Each category involves a different interaction modality with the nurse case manager^45,46^, who will interface between the patient and the corresponding primary care physician (for all patients) and/or specialist (only for severe asthma patients), as delineated in Figure 2.

**Figure 2.**
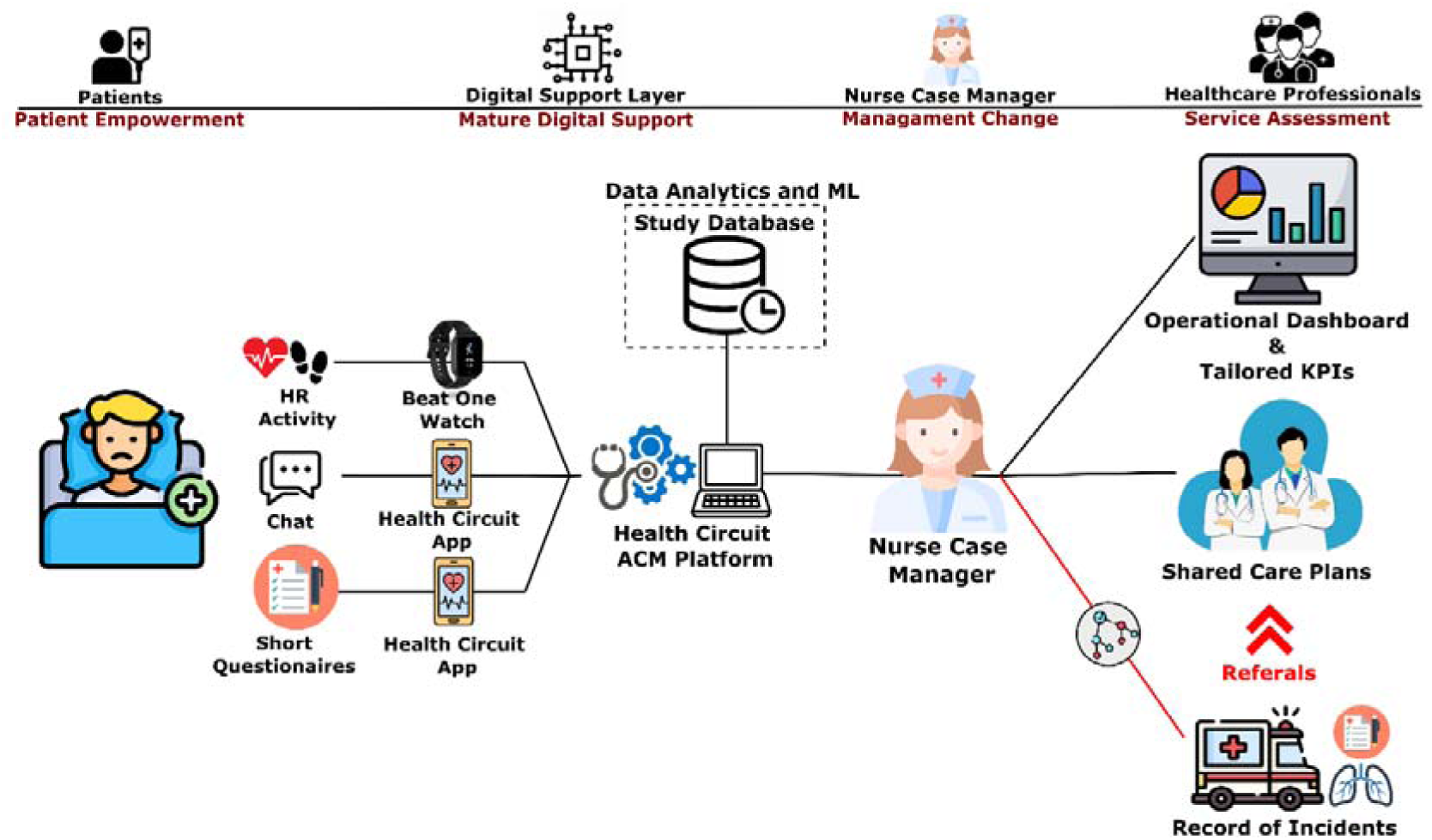
Enhanced patient management scheme. Patients participating in the digitally enabled follow-up will be overseen by a nurse case manager utilising the Health Circuit solution. The Health Circuit patient app offers a trimodal approach to data acquisition: i) Biological data collection throughout the Beat One watch; ii) a chat channel for direct communication with the nurse case manager; and iii) regular administration of health questionnaires to monitor disease status. Collected data through the Health Circuit app will be integrated into the study databases for comprehensive data analytics and predictive modelling. The Health Circuit professionals’ platform, controlled by the nurse case manager, provides enhanced patient management solutions. ML stands for Machine Learning, ACM for Adaptive Case Management and KPI for Key performance indicator.

Briefly, after the baseline assessment, the nurse case manager will undertake a motivational interview with the patient with a twofold purpose. Firstly, it will empower them with self-management strategies. Secondly, it will help them agree on the specificities of a personalised work plan that involves pharmacological and non-pharmacological actions. This health plan will be individually formulated with the patient, the case manager nurse, and the designated physician, whether the primary care physician or a specialised clinician. Such consensus action plan will be video recorded and used during the follow-up for subsequent interactions between the nurse case manager and the patient, utilising the Health Circuit mobile application as a supporting tool.

As depicted in Figure 2, the Health Circuit backend provides the case manager nurse with a dashboard for cohort monitoring and adherence to the agreed health plan, facilitating cohort tracking. The Health Circuit platform enables physiological data collection by synchronising with the Beat One watch and facilitates the periodic administration of health questionnaires for disease monitoring. Additionally, it offers a chat channel for direct communication with the patient. This is accompanied by a standardised cribbage scheme in the form of a decision tree to report health incidents, aiming to detect and intervene early in exacerbation episodes. In the event of reported incidents, the case manager nurse will contact the reference physician to define the course of action and treatment plan. The **Supplementary Material – Appendix 2** describes the specifics of the protocols guiding the interactions between patients and the nurse case manager.

### 2.1 Integrated Care Service Components

As depicted in Figure 1, the development of the current protocol includes achievements in the four main service components executed during 2024; that is: i) Enhanced Lung Function Testing, ii) Continuous monitoring of IAQ, iii) Digital support with an Adaptive Case Management approach, and iv) Predictive modelling for enhanced management of exacerbations. Each component has specificities in design, setup, data analysis, and expected results, described below.

#### 2.1.1. ENHANCED LUNG FUNCTION ASSESSMENT

##### Background and rationale

Using oscillometry to measure the respiratory system resistance and reactance shows a long track of physiological research from its introduction 70 years ago^19^. However, the interest in its clinical application, which is not invasive and does not require active patient cooperation, has steadily grown over the last decade. The 2020 update of the technical standards developed by a European Respiratory Society (ERS) task force of international experts^47^ and recent publications in the field^48–50^ indicate a high potential of the technique in lung function testing. The need for further information on the relationships between oscillometry variables, namely resistance and reactance, and clinical disorders has been acknowledged^49^. Also, better identification of complementarities between this technique and conventional forced spirometry (FS) testing is required. All in all, the establishment of the role of oscillometry in managing respiratory patients, either within the healthcare system, primary and specialised care, or at home through patients’ self-administration of the technique^48–50^, is a partially unmet need.

The current service component has two objectives. First, it investigates the role of oscillometry compared with FS testing in the entire cohort of 200 patients. Second, it explores the usefulness of home-based self-administered oscillometry in the early identification of clinical exacerbations, thus facilitating the early management of acute events.

##### Setup

All cohort participants (n=200) will undertake lung function measurements, oscillometry and FS at baseline and every six months during a two-year follow-up. Therefore, up to five lung function testing measurements will be done in each patient.

A second analysis will include only those individuals in whom the nurse case manager identifies suspicion of an early acute episode through the communication channel or the input data from the patient’s App, as defined during the first PDSA cycle. In such individuals, home-based equipment for self-monitoring oscillometry and short questionnaires to be answered through the App will be provided for daily assessment during a two-week period^51^. The information collected will be used to guide the management of the acute episode. Home visits of health professionals can be scheduled as needed.

Oscillometry and FS measurements will be performed using an Ambulatory Lung Diagnosis System (ALDS) from Lothar Medtech^52^ compliant with ERS technical standards.

##### Data analysis and expected outcomes

The relationships among oscillometry variables, FS testing and clinical symptoms, patients’ traits, and sensor data (i.e., heart rate variability, HRV), indicated in **Table 1**, will be explored.

The primary expected outcomes of this component are that while FS is valid for the characterisation of the patients in terms of diagnosis and disease severity, the results of oscillometry may offer clear advantages for patients’ monitoring in terms of i) logistics of the testing, ii) patient’s tolerance, iii) reliability of the results in non-specialized settings, and, likely, iv) information content. We also hypothesise that the combination of oscillometry, the patient’s reported data, and sensor information will allow us to characterise and support the early management of exacerbation episodes.

#### 2.1.2. CONTINUOUS MONITORING OF INDOOR AIR QUALITY (IAQ)

##### Background and rationale

Understanding the complex links between IAQ and acute health issues in respiratory outpatients remains an unmet need^9–11^. A comprehensive, long-term monitoring cohort holds promise in addressing the hypotheses surrounding IAQ’s impact on respiratory health. It is recognised that poor IAQ in homes may contribute to worsening respiratory symptoms in high-risk outpatients, potentially leading to diminished lung function, impaired quality of life, increased healthcare visits, and even mortality. This hypothesis underscores the potential of specific indoor pollutants and allergens to induce respiratory inflammation, hinder lung function, and alter heart rate variability, necessitating a focused exploration of these pollutants’ physiological effects on respiratory patients within their homes. Characterising IAQ at patients’ residences is, therefore, pivotal in unravelling the mechanisms triggering exacerbations, offering critical insights for tailored interventions aimed at alleviating respiratory issues and enhancing the well-being of these patients.

##### Setup

Exploring the relationship between IAQ and health status encompasses two primary domains: IAQ information and health-related data. The MICA-INBIOT^36^ sensors will be installed at patients’ homes to capture IAQ information, utilising one desktop device per residence. Interconnected via Wi-Fi, these sensors will continuously and autonomously monitor specific IAQ parameters detailed in **Table 1**.

Simultaneously, health-related data will be gathered from diverse sources (**Table 1**): i) Standardized periodic questionnaires covering various dimensions, ii) Patient-generated data through the Health Circuit app (encompassing self-tracked incidences, nurse case manager interactions, Likert scale-based questionnaires), iii) Physiological signals and physical activity via a wrist sensor, and iv) Clinical information encompassing lung function measurements, and registry data sourced from the CHSS. This comprehensive approach holistically allows to capture IAQ dynamics and multifaceted health-related information to elucidate the nuanced relationship between indoor air quality and respiratory patient well-being.

##### Data analysis and expected outcomes

The data analysis will encompass descriptive statistics, correlation, and time series analyses to extract meaningful insights. Descriptive statistics will summarise and present the IAQ parameters monitored by the MICA-INBIOT sensors, providing a clear overview of the indoor air quality profiles across households. Correlation analyses will be instrumental in examining relationships between IAQ parameters and health-related data sources, unveiling potential associations between specific determinants and various health outcomes experienced by respiratory patients. Additionally, time series analyses will offer a dynamic perspective, exploring how IAQ parameters fluctuate over time and their potential impact on acute health issues among respiratory outpatients. Examining trends and patterns in IAQ variations alongside health-related events or exacerbations can reveal potential triggers or associations between changes in indoor air quality and respiratory health outcomes.

#### 2.1.3. DIGITAL SUPPORT WITH AN ADAPTIVE CASE MANAGEMENT (ACM) APPROACH

##### Background and rationale

The pilot study^20^ on community-based management of complex chronic patients with high risk for hospitalisation showed the potential of the ACM approach to enhance the management of unplanned health events. This would lead to less use of health care resources and improved patient empowerment for self-management and perception of continuity of care^53,54^.

The current study has a twofold aim. The first two PDSA cycles will contribute to refining the digital tool’s current functionalities, as shown in Figure 2 and ^20^. Secondly, it will evaluate the potential of a set of short questions answered using a Likert scale, to capture patient-reported outcomes (PROMs) and patient-reported experiences (PREMs).

##### Setup

The analysis of professionals’ (n=20) and patients’ (n=200) usability and acceptability of the digital support (Health Circuit) will be done with the self-administered net promoter score (NPS)^55^, the System Usability Scale (SUS)^56^ and the Nijmegen Continuity questionnaires^57^, respectively. Lastly, an evaluation of the set of short questions, aiming at capturing PROMs and PREMs, will be conducted by comparing the Likert scoring with the results of the corresponding standardised questionnaires, as reported in **Table 1**. Detailed features of the assessments will be defined during the first PDSA cycle.

##### Data analysis and expected outcomes

At the end of the first year, a refined version of Health Circuit properly customized to address the objectives of 2025 is expected to be generated. The corresponding questionnaires will substantiate the usability and acceptability of digital support for patients and health professionals. Finally, validating the set of short questionnaires for capturing PROMs and PREMs should contribute to evaluating the service during Phase II.

#### 2.1.4. PREDICTIVE MODELLING FOR ENHANCED MANAGEMENT OF EXACERBATIONS

##### Background and rationale

The negative impact of exacerbations on quality of life, resource use, disease progression and prognosis has been well documented^12,13^. However, early detection and personalized treatment considering pharmacological and non-pharmacological interventions to effectively prevent emergency room consultations and unplanned hospital admissions is still an unsolved challenge. This is partly due to current post-hoc definitions of exacerbation relying broadly on symptomatic changes. However, this is also explained by inherent heterogeneities in patients’ underlying conditions due to comorbidities.

The innovative study framework proposed herein presents a pioneering approach to this challenge: using machine learning to detect exacerbations early^58^. Leveraging Health Circuit’s patient management paradigm alongside comprehensive patient health assessment holds promise in accurately discerning and characterising exacerbation events.

##### Setup

The nurse case manager will document all instances indicating suspicion of an exacerbation episode detected through the patient’s App input, as well as exacerbation episodes resulting in primary care visits, emergency room visits, or hospitalizations.

##### Data analysis and expected outcomes

Upon concluding Phase I, an analysis of the time-series data encompassing physiological signals, air quality metrics, lung function, patients’ disease perception, and underlying comorbidity will be conducted to characterise these exacerbation occurrences thoroughly. Moreover, diverse modelling strategies to predict exacerbation events will be generated, tested, and compared. The model exhibiting superior performance will be prospectively validated during Phase II. Predictions will not be used for clinical intervention. In addition, explainable AI methods will be employed to assess the model’s covariates (i.e. the mean decrease in accuracy method^59^), determining their predictive efficacy and isolating the critical signals instrumental in early identifying exacerbations (i.e. Anchor method^60^).

By harnessing advanced technological tools and conducting data analyses, this study aims to yield several pivotal outcomes. Initially, the thorough documentation and comprehensive assessment of both suspected and documented exacerbation episodes, alongside a multifaceted evaluation of patient health parameters, seek to refine the precision and accuracy in identifying and characterising exacerbations. We aim to unravel profound insights into exacerbation patterns through an exhaustive analysis of temporal data spanning physiological indicators, air quality metrics, patients’ perceptions of their illness, and comorbidity profiles. This approach is anticipated to expedite early intervention strategies and enable the implementation of more targeted preventive approaches.

Lastly, exploring and comparing diverse predictive modelling methodologies aims to construct robust predictive models for forecasting exacerbation events, catalysing the development of clinical applications and offering decision support tools to aid healthcare providers in efficiently anticipating and managing exacerbations.

### 2.2 Assessment of the Novel Integrated Care Service

As depicted in Figure 1, the achievements in the four main service components executed during 2024 will conform to the final features of the novel integrated care service to be assessed during 2025 concerning the potential for value generation, aiming at preventing unplanned hospital admissions. Moreover, the research will identify key elements ensuring sustainable adoption of the service in a real-world environment and facilitating site transferability of the intervention.

#### Background and rationale

The progress experienced in terms of adoption of integrated care services at the regional level over the last decade^14,61,62^, as well as key lessons learned in recent studies done in AISBE^24^, suggest a high potential to overcome the efficacy-effectiveness gap seen in previous local analyses carried out addressing the prevention of unplanned hospital admissions^63,64^. Moreover, the current research’s two-phase study design ensures our approach’s maturity.

The main objective of the second year is to perform a Quintuple Aim evaluation of the community-based service to prevent unplanned hospital admissions. Also, to assess the process of implementation of the service.

#### Setup

The consolidated version of the service for managing the patients’ cohort (n=200) at the end of Phase I (Figure 1) will be deployed and evaluated over a year in 2025. Health outcomes and patients’ expenditures in the study cohort will be compared with a control group under conventional care selected from the same primary care units.

The control group will comprise AISBE patients with chronic pulmonary diseases during the same timeframe. To establish comparability, study participants will undergo a 1:1 pairing with control outpatients using propensity score matching ^65,66^ and genetic-matching techniques^67^. Two sets of variables will be considered for this matching process to ensure patient comparability. These variables will encompass primary pulmonary diagnoses (e.g., COPD, Asthma, etc.) and baseline characteristics, incorporating factors such as age, gender, socioeconomic status, number of admissions in the preceding year, healthcare costs within the healthcare system during the prior year, and the comorbidity burden assessed by the AMG index.

#### Data analysis and expected outcomes

The Quintuple Aim assessment will include: i) Health outcomes such as number of acute episodes, emergency room consultations, hospital admissions, and mortality, ii) PROMs and PREMs, iii) health professionals’ experience, and iv) direct costs evaluated with a cost consequence analysis. It is worth noting that items involving indirect costs will be identified to assess potential sources of inequity.

The CHSS database will be consulted to compare clinical outcomes between the study participants and a control group. This comprehensive analysis will encompass i) mortality rates; ii) healthcare professional encounters, including primary care visits, specialised outpatient visits, ambulatory mental health centre visits, emergency room visits, planned and unplanned hospital admissions, and admissions to mental health centres; as well as iii) total healthcare expenditure. The latter includes direct healthcare delivery costs, pharmaceutical expenses, and other billable healthcare costs like non-urgent medical transportation, ambulatory rehabilitation, domiciliary oxygen therapy, and dialysis.

The comprehensive assessment planned in the current study should allow the identification of the profile of candidates for the novel service and provide valuable information on its potential for value generation. Moreover, the protocol will evaluate the service adoption process using a simplified CFIR^22^ approach to identify key barriers and facilitators that may guide site transferability. The evaluation of the main items of the CFIR will be quantitatively expressed using Lickert scales. Finally, a profiled dashboard for quality assurance of the service after adoption will be proposed.

## 3. ETHICAL AND REGULATORY ISSUES

The Ethical Committee for Human Research at Hospital Clinic de Barcelona approved the core study protocol of K-Health in Air on June 29, 2023 (HCB/2023/0126). The study design adheres to data minimisation principles, ensuring that only essential data are collected and utilised. The study will be conducted in compliance with the Helsinki Declaration (Stronghold Version, Brazil, October 2013) and in accordance with the protocol and the relevant legal requirements (Biomedical Research Act 14/2007 of July 3).

All patients in the study must sign an informed consent form before any procedure. The participants can withdraw their consent at any time without altering their relationship with their doctor or harming their treatment.

The One Beat watch does not hold medical device certification and will be utilized solely for data collection and exploring potential patterns informing exacerbations, not for decision-making within the study. Any future application derived from this research must ensure that the technology aligns with medical device regulations.

## Data Availability

Data availability does not apply to this article since it describes a protocol rather than presenting study results

## Authors’ Contributions

JR and RGC designed the study and are responsible for overseeing project coordination. JPJ and JF contributed valuable insights into the study design. JR and RF will spearhead the development of the enhanced lung function assessment study. IC, CH, and AGL will take the lead in designing the study on digital support through an adaptive case management approach. RGC and IC will also head the development of predictive modelling for improved management of exacerbations.

MF will provide the MICA sensors on behalf of INBIOT and will offer expertise in IAQ/OAQ assessment jointly with BS. EV will manage the extraction of pertinent data from the CHSS database and provide statistical support.

AGL will fulfil the responsibilities of the nurse case manager as outlined in the protocol, which includes overseeing cohort recruitment and follow-up. The clinical intervention and patient follow-up plans will be collaboratively determined by AGL, EA, NS, MS, ASA, JB, AMI and JR.

AGL, JR, and RGC initially drafted the manuscript, which included writing the text and creating infographics. It was then thoroughly reviewed by IC and CH. All authors have approved the final version of the manuscript and are committed to upholding the integrity and accuracy of the work.

## Disclaimer

Views and opinions expressed are, however, those of the authors only and do not necessarily reflect those of the European Union or the European Health and Digital Executive Agency as granting authority. Neither the European Union nor the granting authority can be held responsible.

## Acronyms

AISBE: Integrated Health District of Barcelona-Esquerra
ACT: Asthma Control Test
AQLQ: Asthma Quality of Life Questionnaire
ALDS: Ambulatory Lung Diagnosis System
AMG: Adjusted Morbidity Groups
CAT: COPD Assessment test
CFIR: Consolidated Framework for Implementation Research
COPD: Chronic Obstructive Pulmonary Disease
FS: Forced Spirometry
HCB: Hospital Clínic de Barcelona
HRV: Heart Rate Variability
ICHOM: International Consortium for Health Outcomes Management
IAQ: Indoor Air Quality
K-HEALTHinAIR: EU Project “Knowledge for improving indoor AIR quality and HEALTH”
mMRC: modified Medical Research Council
NPS: Net Promoter Score
PaRIS: Patient-Reported Indicator Survey
PM: Particulate Matter
PROMIS: Patient-Reported Outcomes Measurement Information System
PROMs: Patient-Reported Outcome Measures
PDSA: Plan-Do-Study-Act cycle
PREMs: Patient-Reported Experience Measures
PS: Propensity Score Matching
SUS: System Usability Scale
SNOT: Sino-Nasal Outcome Test
TAI: Test of Adherence to Inhalers
VOC: Volatile Organic Compound
WHO: World Health Organization

