## Supplementary Material for "Protocol for the Enhanced Management of Multimorbid Patients with Chronic Pulmonary Diseases: Role of Indoor Air Quality"

Alba Gómez et al.

*(On-line supplementary material)*

Table of content

### Appendix 1: Summary of the “Knowledge for improving indoor AIR quality and HEALTH” project (K-HEALTHinAIR – KHiA)

**Project details:**

**Call:** HORIZON-HLTH-2021-ENVHLTH-02

**Type of action:** HORIZON-RIA

**Proposal number:** 101057693

**Proposal acronym:** K-HEALTHinAIR

**Duration (months):** 48

**Proposal title:** Knowledge for improving indoor AIR quality and HEALTH.

**Activity:** HORIZON-HLTH-2021-ENVHLTH-02-02

**Project funding:** (7,984,484 €)

**Project partners:**

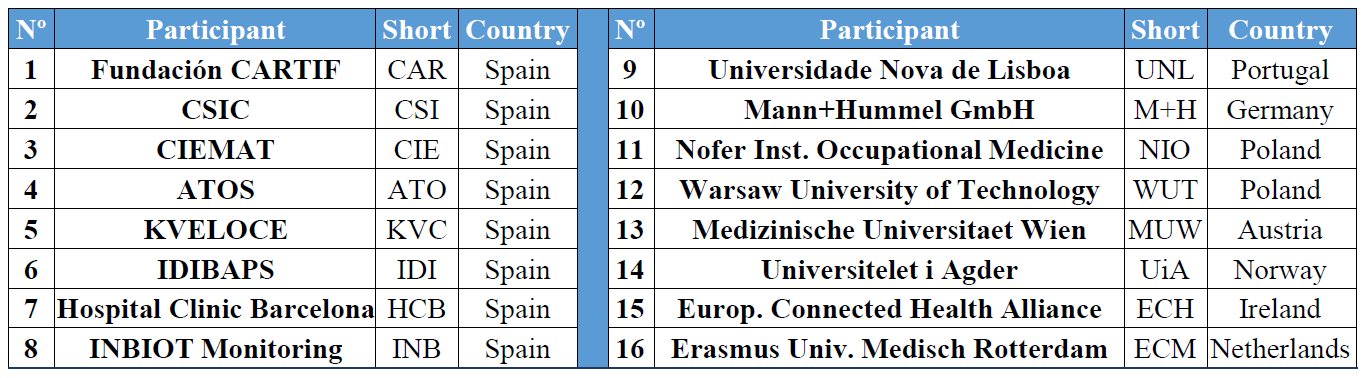

Table S1: Institutions that conform the K-HEALTHinAIR consortium.

**Project summary:**

The K-HEALTHinAIR initiative represents a pioneering interdisciplinary effort to expand our understanding of chemical and biological contaminants in indoor air that influence human health. This project is committed to enhancing the precision and effectiveness of indoor air quality (IAQ) monitoring and improvement strategies. Utilizing groundbreaking artificial intelligence and comprehensive data analysis, the project identifies key determinants, their sources, and the associated health risks of indoor air pollutants. The project outcomes will include a compilation of structured knowledge derived from monitoring, characterization, and research phases, presented in an accessible, open-access format to support public authorities, policymakers, and various stakeholders.

The project undertakes meticulous research, analysing data from authentic environments, health surveillance initiatives, and demographics particularly sensitive to IAQ issues, including high-risk outpatients and specific age groups. It aims to establish comprehensive links between IAQ and its health impacts, focusing on characterizing IAQ across ten European scenarios, especially in areas where IAQ significantly affects large or vulnerable populations, encompassing hospitals, academic institutions, and residential settings, among others. Through five targeted pilot studies (**Figure S1**), this endeavour will produce crucial insights into indoor air quality across ten significant European contexts.

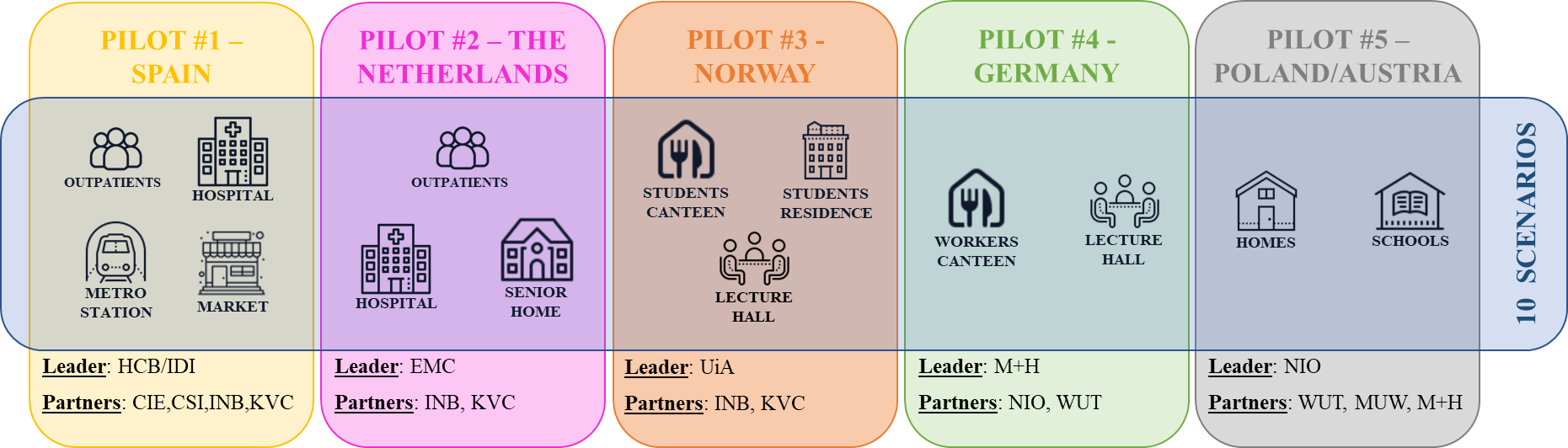

Figure S1: Pilots and scenarios covered within the K-HEALTHinAIR project. The acronyms of the project partners are depicted in Table S1.

**#1 Barcelona pilot**: This comprehensive study encompasses two main objectives: tracking high-risk outpatients with respiratory diseases and evaluating IAQ in hospitals, metro stations, and markets.

**#2 Rotterdam pilot:** Focused on outpatient care, this pilot also examines IAQ in hospitals and communal areas of senior living.

**#3 Norway pilot:** This study, dedicated to assessing IAQ and the health impacts of wooden building materials, investigates a cafeteria, student housing, and a lecture hall.

**#4 Germany pilot:** This pilot aims to pinpoint IAQ determinants in a cafeteria and lecture hall and optimize conditions for occupants.

**#5 Poland/Austria pilot**: This study analyses residential and school settings and seeks IAQ determinants in various domestic environments and educational institutions.

K-HEALTHinAIR integrates IoT devices for continuous and detailed IAQ monitoring, which is essential for addressing health and well-being challenges in enclosed spaces. These sensors are pivotal for real-time air quality assessment and capture data on temperature (Tª), relative humidity (RH), CO2, total volatile organic compound (tVOC) concentration, formaldehyde levels, and various particle sizes (PM1, PM2.5, PM4, and PM10). With a universal communication system, these devices ensure consistent connectivity, supported by a cloud platform for data management and analysis. This setup not only provides comprehensive IAQ insights but also aids in the early detection of health hazards, supporting proactive interventions. Continuous monitoring offers dynamic insights into IAQ fluctuations, facilitating a responsive and adaptable approach to managing the indoor environment.

### Appendix 2: Details of the clinical intervention for enhanced management of multimorbid patients with chronic obstructive pulmonary diseases (i.e. COPD and severe asthma)

The protocol explores the potential positive impact of proactive early management of COPD and asthma exacerbations on patients’ health status and unplanned hospital admissions.

As depicted in **Figure 2** of the main manuscript, the digitally enabled clinical pathway has four main elements: i) the nurse case manager, ii) the pro-active patient, iii) the reference doctor, and iv) the patient’s self-captured information, including symptoms, self-administered short questionnaires, and real-time monitoring of physiological variables, such as heart rate variability (HRV) and activity levels.

**Personalized patient’s action plan** - The initial step involves a motivational interview with the patient conducted by the nurse case manager. This interview serves two main purposes. Firstly, it aims to empower the patient for self-management of their condition. Secondly, it facilitates the collaborative development of a patient’s tailored care plan, with a holistic approach, agreed upon by the patient, the nurse case manager, and the reference doctor. This action plan forms the basis for the patient’s periodic follow-up with the nurse case manager, with most interactions taking place through the chat.

**Management of exacerbations** – In the event of a worsening of clinical conditions the patient would initiate contact with the nurse case manager through the chat. This contact triggers the process of identifying a potential exacerbation, based on the patient's reported symptoms, scoring of a short questionnaire (CAT for COPD and ACT for asthma patients), and the clinical judgement of the nurse.

The suspicion of an exacerbation will activate the following actions: i) close follow-up of the patient, remote or face-to-face, by the nurse case manager, ii) daily monitoring of the patient with self-administration of a short questionnaire (CAT or ACT depending upon the main diagnose), as well as home-based measurements of oscillometry, and iii) adjustment of the therapeutic plan in agreement with the reference doctor. Such action plan will be maintained during one or two weeks depending upon the patient’s evolution.

As described in the main manuscript (**Figure 1**), the setting must facilitate the development, refinement, and evaluation of a machine learning-based model (including symptoms, PROMs, oscillometry, and physiological data from wearable devices) to predict the onset of an exacerbation, as well as the severity of the acute episodes. The objective of such predictive modelling is to provide support to health professionals for clinical decision making facilitating early identification and management of exacerbations.

The development, refinement and evaluation of the predictive modelling tool will be done through the co-design process depicted in **Figure 1** of the main manuscript and detailed in the **Appendix 3**. This co-design process, encompassing several dimensions of the innovative integrated care service for enhanced management of the patients, is conceived as four sequential Plan-Do-Study-Act (PDSA) cycles of six-month duration each.

Specifically for the development, and assessment, of the predictive modelling tool, the objectives of each PDSA cycle are as follows: i) Optimization of clinical applicability of data capturing to facilitate the construction of the predictive modelling - PDSA 1, January to June 2024; ii) Elaboration of the machine learning-based model – PDSA 2, July-December 2024; iii) Model training and refinement – PDSA 3, January-June 2025, and iv) Clinical testing of the model and design of quality assurance following the APPRAISE-AI recommendations^1^ – PDSA 4, July-December 2025.

### Appendix 3: Plan-Do-Study-Act (PDSA) cycles template

The main manuscript describes a two-phase prospective follow-up study involving 200 outpatients scheduled from 1st January 2024 to 31st December 2025. The first phase, spanning the entire year of 2024, aims to consolidate the characteristics of the new service by the year's end, focusing on the detailed implementation of its four primary components. The second phase, covering the entirety of 2025, aims to accomplish three key objectives: assessing the service outcomes, identifying patient profiles suitable for preventive interventions, and evaluating the service implementation process.

The integration of the service's components as outlined in the protocol will be facilitated through four sequential six-month PDSA (Plan-Do-Study-Act)^2^ cycles. The PDSA cycle represents a methodological framework employed in various domains, notably healthcare, education, and organizational management, to facilitate empirical, iterative testing and refinement of changes implemented within processes or products. This framework is instrumental in the empirical underpinning of evidence-based practice, allowing for the methodical evaluation and enhancement of procedures or systems. The four phases of the PDSA cycle are elucidated below:

**Plan:** This phase entails the identification of a target domain for enhancement and formulation of a strategic approach to effectuate modifications. It involves delineating objectives, formulating hypotheses regarding the potential outcomes of the proposed changes, and comprehensively planning the implementation process. Baseline data acquisition is imperative to establish a reference point for subsequent comparative analysis.

**Do:** During this phase, the devised plan is executed on a pilot scale to test the proposed changes. It necessitates meticulous documentation of any encountered challenges or unanticipated observations. Concurrent data collection is paramount to facilitate a subsequent analytical evaluation of the intervention's impact.

**Study:** This phase involves the analytical assessment of the data garnered in the Do phase, juxtaposing the actual outcomes against the anticipated results to ascertain the efficacy of the intervention. This analytical process is critical in deducing the success level of the implemented change and in gleaning insights from the empirical findings.

**Act:** Predicated on the analytical insights gleaned from the Study phase, this step involves making informed decisions regarding the future course of action. Should the intervention prove efficacious, scaling it up for broader implementation might be warranted. Conversely, if the outcomes do not align with the desired objectives, the insights acquired can inform the planning of subsequent PDSA cycles, thereby instigating a refined approach to the intervention.

This iterative, cyclical approach epitomizes the character of continuous improvement and incremental learning, fostering a systematic methodology for enhancing processes or products through empirical testing and adaptive learning. Within this project, the anticipated outcomes after the completion of the PDSA cycles include the following: during **PDSA-1**, selection of clinically applicable patients’ data capture; in **PDSA-2**, the finalization of the service design; throughout **PDSA-3**, an initial evaluation of the predictive models for early detection of exacerbations to support AI-enhanced patient management; and in **PDSA-4,** a thorough evaluation of the new service's effectiveness.

The PDSA methodology has been extensively utilized in prior evaluations^3^, resulting in generating tailored templates for each cycle segment, as delineated below. These personalized templates are customized to streamline and standardize the application of the PDSA framework in healthcare. See below, as an example, the description of first step (Plan) of the PDSA cycle 1. As reported in the main document, the PDSA cycle 1 is currently running from 1^st^ January (M1) to 30^th^ June 2024 (M6).

**Plan (cycle 1)**

| **Task** | Clinical Applicability of Data Capture: Definition of the protocols and feasibility analyses | | | | | | | |
| --- | --- | --- | --- | --- | --- | --- | --- | --- |
| **Activities** | **Actions** | **Actors** | **Timeline**  **(Month)** | **KPIs measure (data collection)** | | | | |
|  |  |  |  | **KPI** | **Who** | **When** | **How** | **Target** |
| Lung Function Testing:  Oscillometry  *(Quality Assurance plan*  *to be defined in the protocol– QA plan)* | Baseline  Measurements | 1.K-Health team  2.Lung Function Lab  3.Biophysics Unit | M1-M3 | Defined in the  QA plan | Team 1 | M6 | Defined in the  QA plan | Defined in the  QA plan |
|  | Plan home-based  measurements |  |  |  |  |  |  |  |
| Predictive Modelling  of exacerbations  *(Quality Assurance plan*  *to be defined in the protocol – QA plan)* | Availability of IAQ data | 1.K-Health team  2. CSIC  3.Biophysics Unit | M1-M3 | Defined in the  QA plan | Team 1 | M4 | Defined in the  QA plan | Defined in the  QA plan |
|  | Availability of clinical data |  |  |  |  |  |  |  |
|  | Availability of wrist  sensor data |  |  |  |  |  |  |  |
| Digital support for patients’ data capturing  *(Quality Assurance plan*  *to be defined in the protocol – QA plan)* | Robustness/Usability | 1.K-Health team  2. Health Circuit S.L. | M1-M3 | Defined in the  QA plan | Team 2 | M4 | Defined in the  QA plan | Defined in the  QA plan |
|  | Questionnaires |  |  |  |  |  |  |  |
|  | Chat |  |  |  |  |  |  |  |
|  | Wrist sensors |  |  |  |  |  |  |  |

*K-Health team stands for the core team of researchers at IDIBAPS (RGC, AGL, and IC); Lung Function Lab corresponds to the unit at Hospital Clinic de Barcelona (HCB), led by EA; Biophysics and Bioengineering Unit at the School of Medicine and Health Sciences of the University of Barcelona, led by RF; and Health Circuit S.L. is a start-up company from IDIBAPS-HCB, IC is the CEO.*

**Do**

| **Cycle number (1)** | Clinical Applicability of Data Capture: Definition of the protocols and feasibility analyses | |
| --- | --- | --- |
| **Activity** | **KPI** | **Actual value** |
| Lung Function Testing:  Oscillometry |  |  |
| Predictive Modelling  of exacerbations |  |  |
| Digital support for  patients’ data capturing |  |  |

| **QUESTIONS** | **ANSWERS** |
| --- | --- |
| **What was actually implemented? Any deviation from the planned actions** |  |
| **Problems? Unexpected findings? Please describe** |  |

| **IMPLEMENTATION PROGRESS** | | | |
| --- | --- | --- | --- |
| **0-25%** | **25-50%** | **50-75%** | **75-100%** |

**Study**

| **Cycle number (1)** | | Clinical Applicability of Data Capture: Definition of the protocols and feasibility analyses | | | | |
| --- | --- | --- | --- | --- | --- | --- |
| **Activity** | **KPI** | **Target value** | **Actual value** | **Reasons for the deviations** | **Mitigation actions implemented** | **Impact of mitigation actions** |
| Lung Function Testing:  Oscillometry |  |  |  |  |  |  |
| Predictive Modelling  of exacerbations |  |  |  |  |  |  |
| Digital support for  patients’ data capturing |  |  |  |  |  |  |

**Act**

| **Cycle number (1)** | Clinical Applicability of Data Capture: Definition of the protocols and feasibility analyses | | |
| --- | --- | --- | --- |
| **Activity** | **Maintain** | **Adapt** | **Abandon** |
| Lung Function Testing:  Oscillometry |  |  |  |
| Predictive Modelling  of exacerbations |  |  |  |
| Digital support for  patients’ data capturing |  |  |  |

| **QUESTIONS** | **ANSWERS** |
| --- | --- |
| **Any new proposed action for the future?** |  |
